# Epidemiology and Clinical Impact of Clonal Hematopoiesis in People with HIV

**DOI:** 10.64898/2025.12.17.25342413

**Authors:** Valeriia Timonina, Konstantin Popadin, Mariam Ait Oumelloul, Alexandra Calmy, Matthias Cavassini, Gioele Capoferri, Huldrych F. Günthard, Laura N. Walti, Patrick Schmid, Philip E. Tarr, Christian W. Thorball, Alex G. Bick, Jacques Fellay, the Swiss HIV Cohort Study (SHCS)

## Abstract

Clonal hematopoiesis (CH), defined by the expansion of hematopoietic cells with somatic mutations in leukemogenic genes (CH of indeterminate potential (CHIP)) or with mosaic chromosomal alterations (mCAs), is associated with aging and adverse health outcomes in the general population. CHIP prevalence has been shown to be higher in People with HIV (PWH) than in controls. However, the full spectrum, prevalence, and clinical consequences of CH in PWH remain incompletely understood. Here, we provide a comprehensive assessment of CHIP and mCAs in a large sample of PWH (N∼2,500) from the Swiss HIV Cohort Study. Using high-depth targeted sequencing of CHIP genes and genome-wide genotyping to call mCAs, we quantified the prevalence and clone size of both types of CH. CHIP (found in 25% of individuals) and mCAs (found in 16% of individuals) were found to be common, positively correlated with age, often co-occurring (OR=1.7, p=0.02 for autosomal mCAs), and associated with various clinical outcomes, including all-cause mortality (HR=1.3, p=0.02 for CHIP) and hematologic malignancies (HR=9.4, p=0.01 for the effect of CHIP on the risk of myeloid cancer; HR>10, p<0.001 for the effect of co-occurring CHIP and mCAs on the risk of lymphoid cancer). We also observed associations of CH with several proxies of inflammatory status (CD4:CD8 ratio, HIV viral load, late initiation of antiretroviral therapy, and toxicity of antiretroviral drugs), highlighting a potential interaction between CH and chronic immune activation.

## INTRODUCTION

Clonal hematopoiesis (CH), the expansion of blood cell clones carrying somatic genetic variants, is associated with aging and adverse health outcomes in the general population^1,2^. Two forms of CH are recognized: clonal hematopoiesis of indeterminate potential (CHIP), which is caused by single nucleotide driver mutations in leukemia-associated genes such as *DNMT3A*, *TET2*, and *ASXL1*, occurring at variant-allele frequencies (VAF) ≥2% in individuals without hematological malignancies^3,4^; and mosaic chromosomal alterations (mCAs), which comprise gains, losses, or copy-neutral loss-of-heterozygosity affecting large chromosomal segments, including sex-chromosome loss (mosaic loss of the Y chromosome (mLOY) in men and mosaic loss of the X chromosome (mLOX) in women) as well as autosomal aneuploidies^5–7^. In the general population, the prevalence of CHIP and mCAs increases with age, and both are associated with cardiovascular disease, hematological malignancies, and all-cause mortality^5–8^.

CHIP contributes to systemic inflammation by altering the function of innate immune cells, thereby linking CH and chronic inflammation to aging and age-related diseases^9,10^. Mechanistically, mutations in CHIP-associated genes like *TET2* have been shown to increase the expression of inflammatory mediators in innate immune cells, reinforcing a cycle of inflammation and clonal expansion that contributes to hematopoietic and systemic aging^9^. This synergistic interaction between CHIP and inflammation in promoting aging-related pathologies^11,12^ has been observed for cardiovascular disease^13^, chronic kidney disease^14^, chronic liver disease^15^, and osteoporosis^12,16^.

On the other hand, mCAs have not been directly linked to increased inflammation. However, they are associated with an increased risk of infections, including sepsis and COVID-19 hospitalization, suggesting they may contribute to the immune system dysregulation^17^.

Most people with HIV (PWH) achieve long-term viral suppression and maintain normal immune function in the setting of effective antiretroviral therapy (ART), and the median age of many national cohorts of PWH now exceeds 50 years. Nonetheless, PWH often show evidence of accelerated aging, with earlier manifestation of aging-related diseases compared to the general population^18^. In addition, non-AIDS cancers and cardiovascular disease have emerged as the major causes of death in this population^19^. This phenomenon might be driven by low-level, chronic immune activation due to residual viremia or gut translocation of bacterial products, by ART toxicity, and by epidemiological and lifestyle factors^20,21^. CHIP prevalence has been shown to be increased in PWH^22–24^. As an example, we previously observed a higher prevalence of CHIP (OR: 1.77, 95% CI: 1.33–2.21, p = 0.02) in Swiss HIV Cohort Study (SHCS) participants compared to age-matched controls^22^. Conversely, there is currently no conclusive evidence of an enrichment of mCAs in PWH^25^.

Here, we provide a comprehensive assessment of CHIP and mCAs in a large sample of PWH. Using high-depth targeted sequencing of CHIP genes and genome-wide genotyping array-based detection of mCAs, we quantified the prevalence and clone size of CHIP and mCAs. We looked for the association of CH with demographic and clinical factors, highlighting potential causes and consequences of CH in PWH.

## METHODS

### Study population

Our study population includes PWH enrolled in the SHCS, a multicentre, open-label, longitudinal, prospective cohort study in Switzerland, which has recruited more than 22,000 PWH since 1988.

The SHCS was approved by the ethics committees of the participating institutions, and written informed consent was obtained from all participants.

For CHIP screening, we included all SHCS participants >= 65 years old who had a sample available for DNA analysis (i.e., DNA extracted from peripheral blood mononuclear cells (PBMCs), or PBMCs available for DNA extraction). If the selected participant had a sample with extracted DNA within 5 years of the last blood sampling, the available sample was used for analysis, lowering the effective minimum age at screening to 60 years. We excluded samples with insufficient DNA or sequencing quality, and patients diagnosed with hematological malignancies before CH screening.

For mCAs detection, we included all SHCS participants previously genotyped using the Global Screening Array-Multiple Disease (GSA-MD) v3 genotyping array from Illumina, who also had information about the date of the sample used for genotyping, excluding individuals diagnosed with hematological malignancies before diagnosis.

### Targeted CHIP sequencing

Targeted, high-coverage DNA sequencing for CHIP detection was performed at the Vanderbilt Technologies for Advanced Genomics (VANTAGE) core facility, using Twist Biosciences’ hybrid capture technology to sequence 24 genes known to be involved in CHIP (the so-called “CHIP genes”) at a depth of greater than 500x to ensure the accurate identification of low-frequency somatic variants^26^. The genes include: *ASXL1, ASXL2, BRCC3, CBL, DNMT3A, ETNK1, GNAS, GNB1, IDH1, IDH2, JAK2, KIT, KRAS, MPL, NRAS, PPM1D, SETBP1, SF3B1, SRSF2, TET2, TP53, U2AF1, ZBTB33,* and *ZNF318*. In total, these genes (or curated portions of these genes that might harbor known CHIP variants) encompass ∼45kb of DNA. The characterization of the 24 genes or portions of genes included on this panel allows the identification of >95% of known CHIP mutations^22,27^.

### Identification of CHIP variants

After the mapping of sequencing reads using BWA-MEM to the GRCh38 reference genome assembly, we used GATK MuTect2 and manual curation to detect the presence of CHIP in each sequenced individual, as described previously^22,27^. We retained the variants with sequencing depth of at least 20, with a minimum of 3 reads mapped to the alternative allele, and at least one forward and reverse sequencing read supporting the variant. Finally, we only kept variants with variant allele frequency (VAF) higher than or equal to 2%.

### Identification of mosaic chromosomal alterations

We identified mCAs from genotyping array data using the MoChA pipeline (https://github.com/freeseek/mocha), following the recommended workflow^5,6^. Briefly, genotype array intensities were first processed using Illumina’s GenomeStudio to generate B-allele frequency (BAF) and log R ratio (LRR) values. Genotype phasing was performed using SHAPEIT4^28^. The phased genotypes and intensity data were then analyzed with MoChA to detect mosaic copy number and copy-neutral loss of heterozygosity (CN-LOH) events, leveraging deviations in phased BAF and LRR. MoChA was run with default settings for autosomes and sex chromosomes, and high-confidence mCA events were retained after standard quality control filters, removing the samples with a call rate less than 97%, phased BAF auto-correlation across consecutive phased heterozygous sites of more than 0.03. For the remaining samples, we removed variants classified as germline copy number polymorphisms. Finally, we applied combined filters on BAF deviation (bdev), relative coverage (rel_cov), and event length: (1) rel_cov < 2.1 or bdev < 0.05, (2) length > 500 kb with bdev < 0.1 and rel_cov < 2.5, or (3) length > 5 Mb with bdev < 0.15. We used the GRCh38 genome reference.

### Statistical analyses

We searched for associations with demographic and clinical factors for the most common types of CH: CHIP status (yes/no); number of CHIP variants; presence/absence of variants in the most common CHIP genes (*DNMT3A, TET2, ASXL1, PPM1D,* and *TP53* for all analyses); any mCAs (yes/no); number of mCAs; autosomal mCAs (yes/no); mLOY in males (yes/no) and mLOX in females (yes/no).

We used Fisher’s exact test to investigate the co-occurrence of CHIP and mCAs. All statistical analyses were performed using R statistical software v.4.5.0.

### Associations of CH with demographic and clinical factors

We first conducted univariate association analyses using logistic regression, adjusting for age, searching for associations between the various types of CH described above and the following variables: sex, genetic ancestry (European vs others), ever-smoking status, injection drug use (IDU), estimated duration of HIV infection, source of infection, number of drugs used for ART, number of ART drugs normalized to the duration of ART, duration of ART (years), duration of untreated infection (years), duration of HIV-infection on ART (%), duration of untreated infection (%), time with detectable viral load (VL) during treatment to total duration of treatment (%), HIV RNA load in copies/ml (mean of all values within one year of CH screening), ART started before year of 2000 (as proxy of treatment toxicity), coinfection with hepatitis B virus (HBV) or hepatitis C virus (HCV), and nadir of CD4 T cells.

Secondly, we included all variables that were associated with any of the CH types with a p-value < 0.1 in multiple regression analyses. We pruned highly collinear variables (Spearman’s rho > 0.6), retaining the one with the strongest effect on CH. With the remaining set of variables, we performed backward selection based on AIC values, using the *stepAIC* function in the *MASS* R package.

Additionally, we fitted a mixed linear model of all longitudinal measurements of HIV RNA load (copies/ml) versus CHIP/mCAs status, adjusting for age, time since ART start, sex, ethnicity, smoking, and IVDU, with a random intercept for each individual.

### Association of CH with blood counts

We investigated associations of CHIP/mCAs status with the following blood parameters: CD3, CD4, and CD8 T lymphocyte counts, total lymphocyte count, leukocyte count [cells per μl], platelet count [10^9/l], hemoglobin [g/dl], lymphocytes as proportion of leucocytes, CD3, CD4, and CD8 T cells as proportions of lymphocytes, and CD4:CD8 ratio.

We fitted a linear model of the mean of blood parameters within one year of CHIP screening or mCAs genotyping versus CHIP/mCAs status, adjusting for age, duration of ART in years, sex, genetic ethnicity, smoking, and IDU.

We also fitted a mixed linear model of all blood parameter measurements versus CHIP/mCAs status, adjusting for age, time since ART initiation, sex, genetic ethnicity, smoking, and IDU, with a random intercept for each individual.

### Association of CH with all-cause mortality and age-related diseases

We conducted a survival analysis using the Cox proportional hazard model (*coxph* function in R), examining the effects of CHIP and mCAs on all-cause mortality and the following age-related diseases: cardiovascular disease (CVD), chronic kidney disease (CKD), liver fibrosis, osteoporosis, hematological malignancies, and solid tumors.

A specific set of covariates (See Outcome definition and covariates in Supplemental Table 1) was used for each model to adjust for known common and HIV-related risk factors^29–33^. For each clinical outcome, we excluded participants who had an event before CHIP/mCAs screening.

### Manuscript preparation

During the preparation of this manuscript, the authors utilized GPT-4 to improve writing and readability.

## RESULTS

### Epidemiology of CHIP and mCAs

Of the 1,693 SHCS participants who met our selection criteria for CHIP screening, we further excluded 57 samples due to insufficient DNA or sequencing quality and 7 participants diagnosed with hematological malignancies before CHIP screening, resulting in a study population of 1,629 individuals.

We observed that 25% of study participants (N=411 out of 1,629) carried at least one CHIP variant. In total, 512 variants were found across 13 genes, with VAF ranging from 2% to 44% (mean = 6.3%) (Figure 1A-C). Increasing the VAF threshold to 5% and 10% decreased the number of CHIP carriers to 10% (N=163 individuals) and 3.7% (N=60 individuals), respectively. The majority (80%, N=335) of CHIP carriers had only one CHIP variant. The most commonly observed CHIP genes were *DNMT3A* (44%), *TET2* (29%), and *ASXL1* (9%). All CHIP variants detected in the current study are listed in Supplemental Table 2.

**Figure 1.**
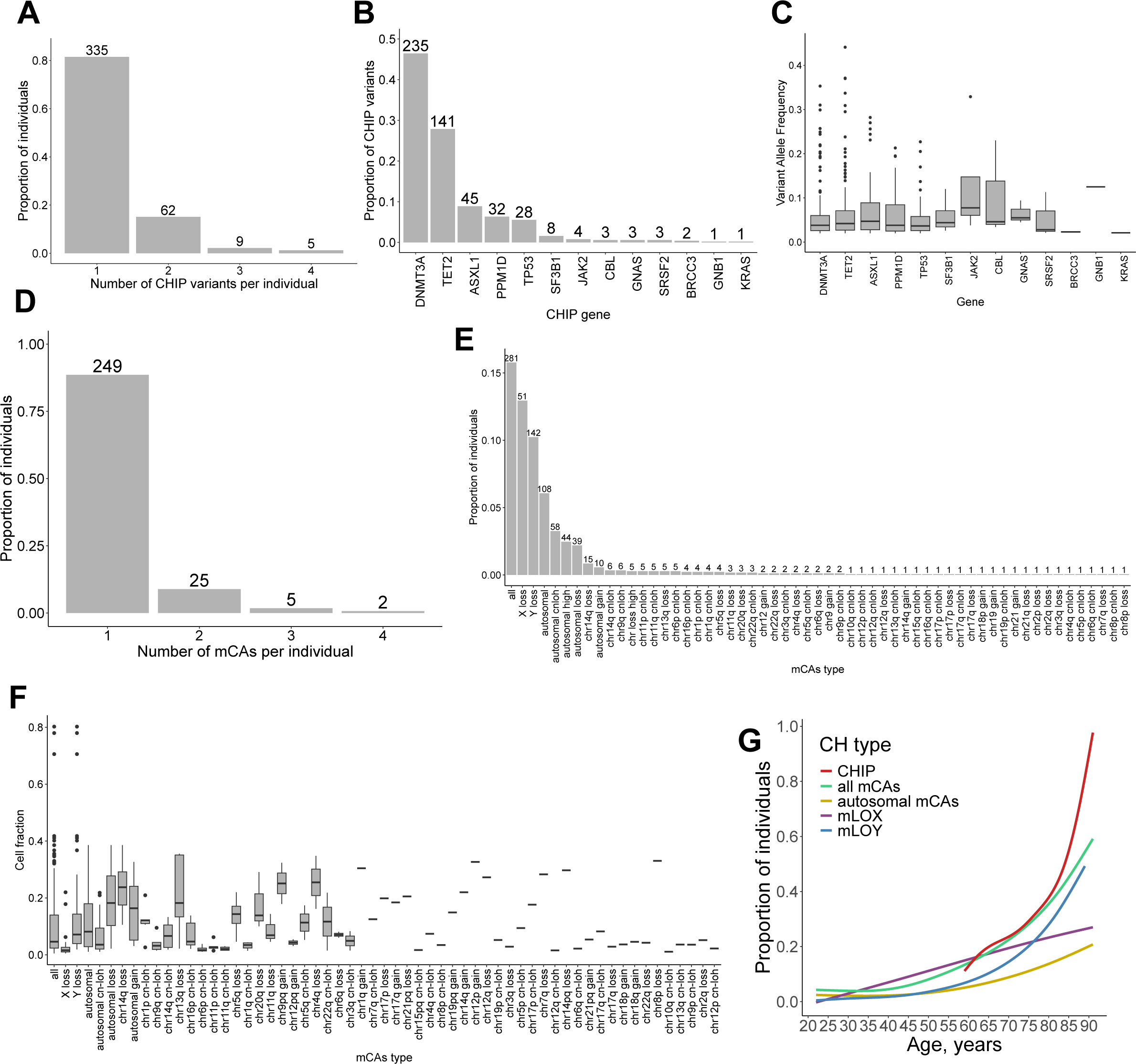
The landscape of clonal hematopoiesis (CH) in study participants. A - Number of CHIP variants per individual among CHIP carriers. Most participants carried a single mutation. B - Distribution of CHIP variants across the 24 targeted genes, with *DNMT3A*, *TET2*, and *ASXL1* being the most frequent. C - Variant Allele Frequency of CHIP variants by gene. All values are above 0.02, which is a conventional Variant Allele Frequency threshold for CHIP definition. D - Number of mCAs per individual among mCAs carriers. E - Prevalence of different mCAs types. The proportions of individuals with X loss and Y loss were calculated for females and males, respectively. F - Estimated fraction of cells carrying different types of mCAs. G - Age-dependent prevalence curves for different CH categories. On A, B, D, and E, the numbers above the bars indicate the number of individuals. On E-G, “all” represents the presence of any mCA.

We detected mCAs in all SHCS participants genotyped with the GSA-MD v3 genotyping array from Illumina, who also had information about the date of the sample used for genotyping (N=1,789). We excluded 7 individuals diagnosed with hematological malignancies before the genotyping date, resulting in a study population of 1,782 individuals.

16% of individuals (n=281 out of 1,782) carried at least one mCA: autosomal mCAs in 6% (N=106), mLOY in 10% of males (N=141), and mLOX in 13% of females (N=51). Among autosomal mCAs, CN-LOH was the most common type (54%, N=58), followed by losses (36%, N=39) and gains (9%, N=10). The majority (89%, N=249) of mCAs carriers had only one mCA. The estimated fraction of cells with mCAs ranged from 0.5% to 80% (mean = 9.1%), with different distributions for the various mCA types (Figure 1D-F). All mCAS variants detected in the current study are listed in Supplemental Table 3.

### Co-occurrence of CHIP and mCAs

A total of 928 individuals screened for both CHIP and mCAs were available to analyze the co-occurrence of different CH types. In this subset of study participants, we observed a co-occurrence of CHIP and mCAs in 96 individuals (OR for enrichment=1.3, p-value=0.08), and an increased co-occurrence of CHIP and autosomal mCAs (OR=1.7, p-value=0.02, Figure 2A). The associations were also observed for general CHIP status and some specific CHIP genes with any mCAs and autosomal mCAs, but not with mLOY or mLOX (Figure 2A and 2B). Among individuals with co-occurring CHIP variants and mCAs, only seven cases involved events affecting overlapping genomic regions—that is, where the CHIP variant was located within the chromosomal segment affected by the mCA (Supplemental Table 4). Across all co-occurring cases, the fraction of cells harboring the CHIP variant and the mCA showed a modest positive correlation (Spearman’s rho = 0.18, p-value = 0.04). This correlation was substantially stronger in the subset with overlapping genomic regions (Spearman’s rho = 0.79, p-value = 0.03), suggesting potential clonal co-localization.

**Figure 2.**
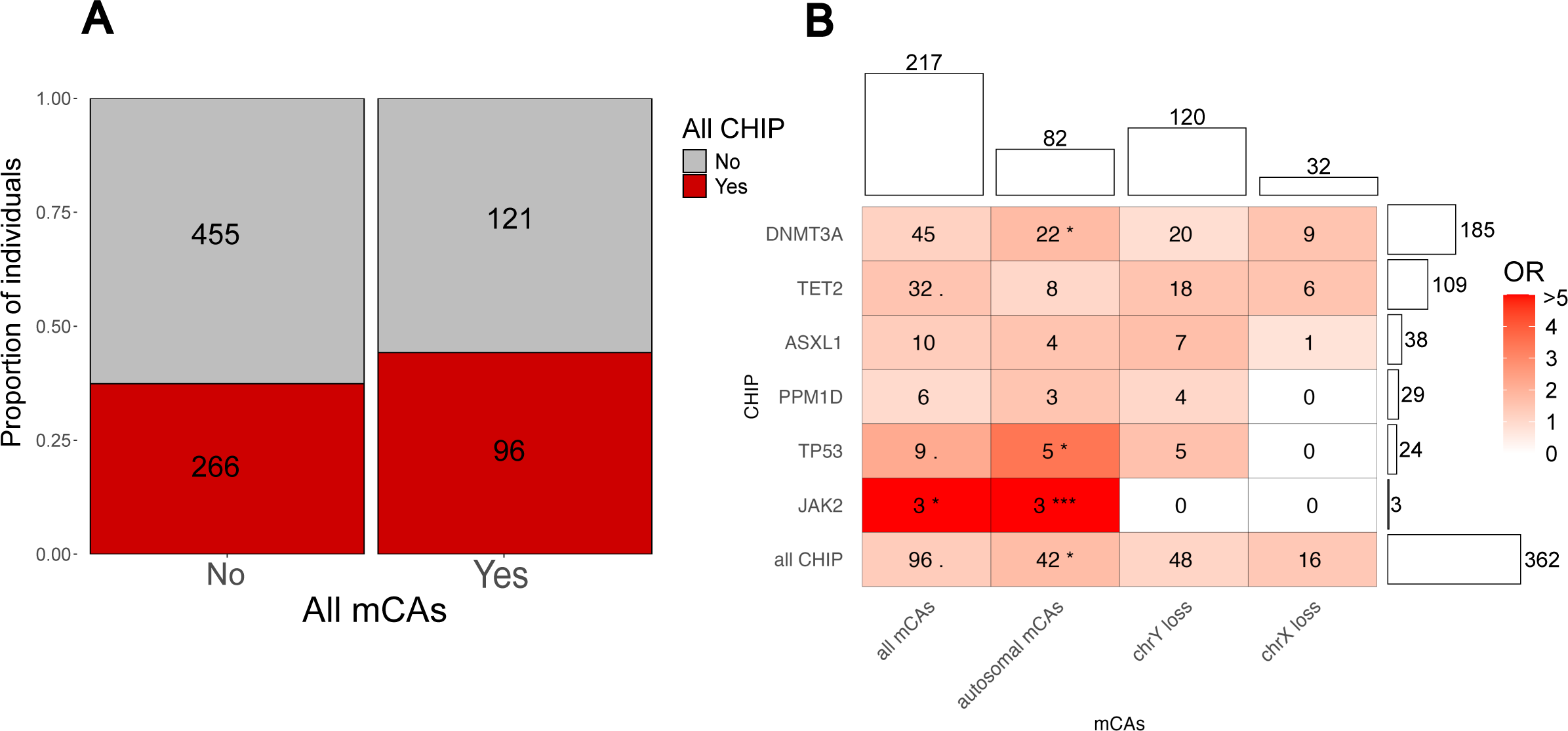
Co-occurrence of CHIP and mCAs in a subset of 928 study participants. A - The co-occurrence of CHIP variants in all targeted genes (y-axis) and all types of mCAs (x-axis). B - Heatmap of co-occurrence between different types of CHIP (y-axis) and mCAs (x-axis). Numbers in each cell represent the number of individuals carrying both mCAs and CHIP. Color represents the OR from Fisher’s exact test. Bars above and on the right show the number of individuals carrying a specific CH. Different p-value thresholds represent the strength of the association:. p-value<0.1; * p-value<0.05; ** p-value < 0.01; *** p-value<0.001.

### Associations with demographic and HIV-related variables

The proportion of individuals with CHIP and mCAs increased sharply with age (Figure 1G). mCAs, but not CHIP, were generally more common in males and individuals of European ancestry. Smoking and IDU were not associated with any of the CH types in univariate analyses. The individuals who carried both CHIP and mCAs were older than those who carried only one CH type. No other demographic or clinical factor was associated with the co-occurrence of CHIP and mCAs (Table 1).

**Table 1.**
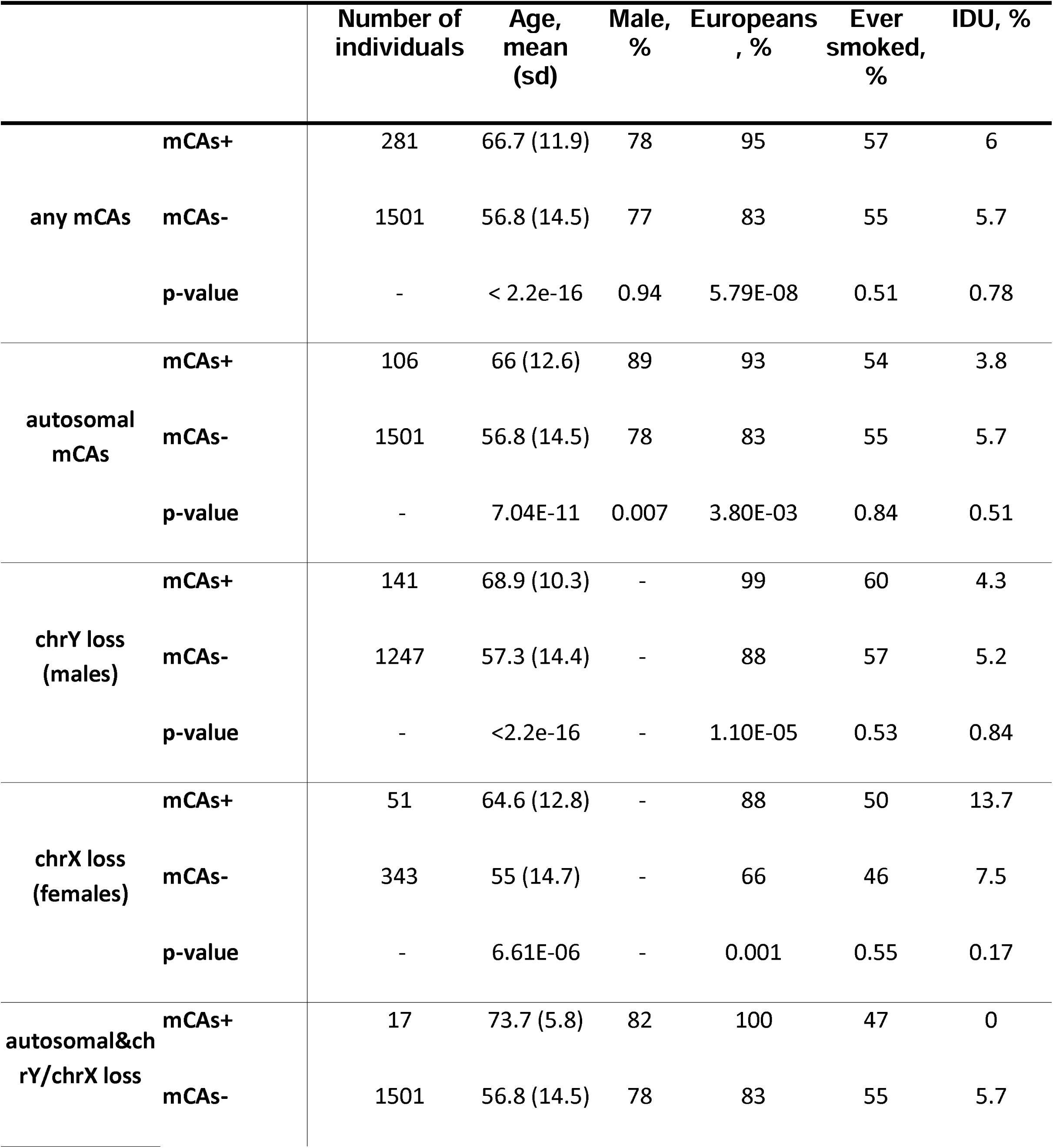
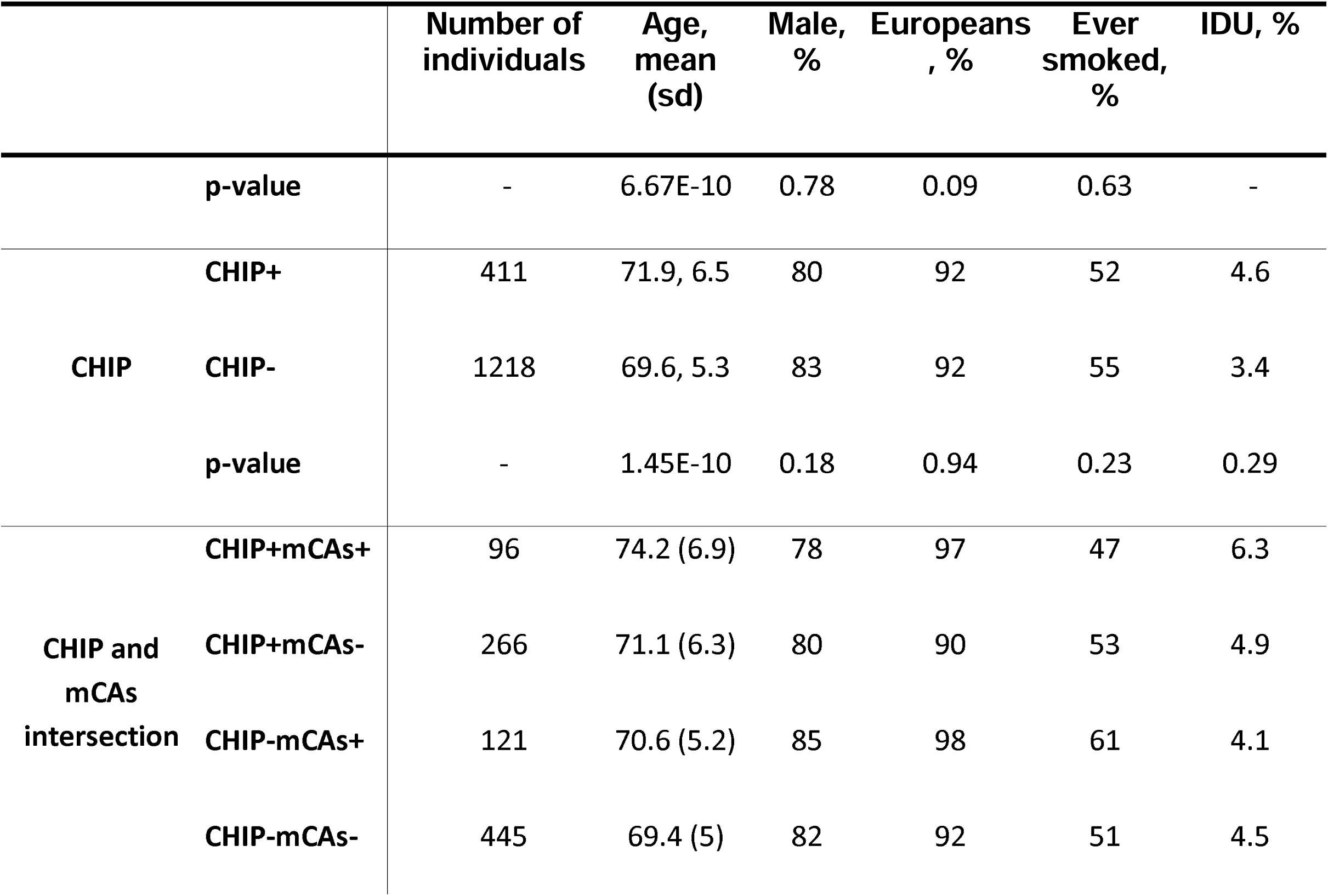
Demographic description of SHCS cohort sample used in this study.

We then searched for associations of the various types of CH with demographic and HIV-related factors in a multivariate analysis (see Methods for the full list of tested factors). Variables with suggestive associations in the univariate analysis (p-value < 0.1, see Supplemental Tables 5-7; age was included as a covariate in all univariate analyses) were included in a multiple regression analysis with backward selection. The final models with the coefficients of the selected variables are presented in Figure 3A. Age was by far the strongest factor, strongly associated with all CH types. Among HIV-related factors, the strongest positive association was observed for IDU status, and the strongest negative association was observed for the proportion of time on ART relative to total infection duration (Figure 3A, Supplemental Table 8).

**Figure 3.**
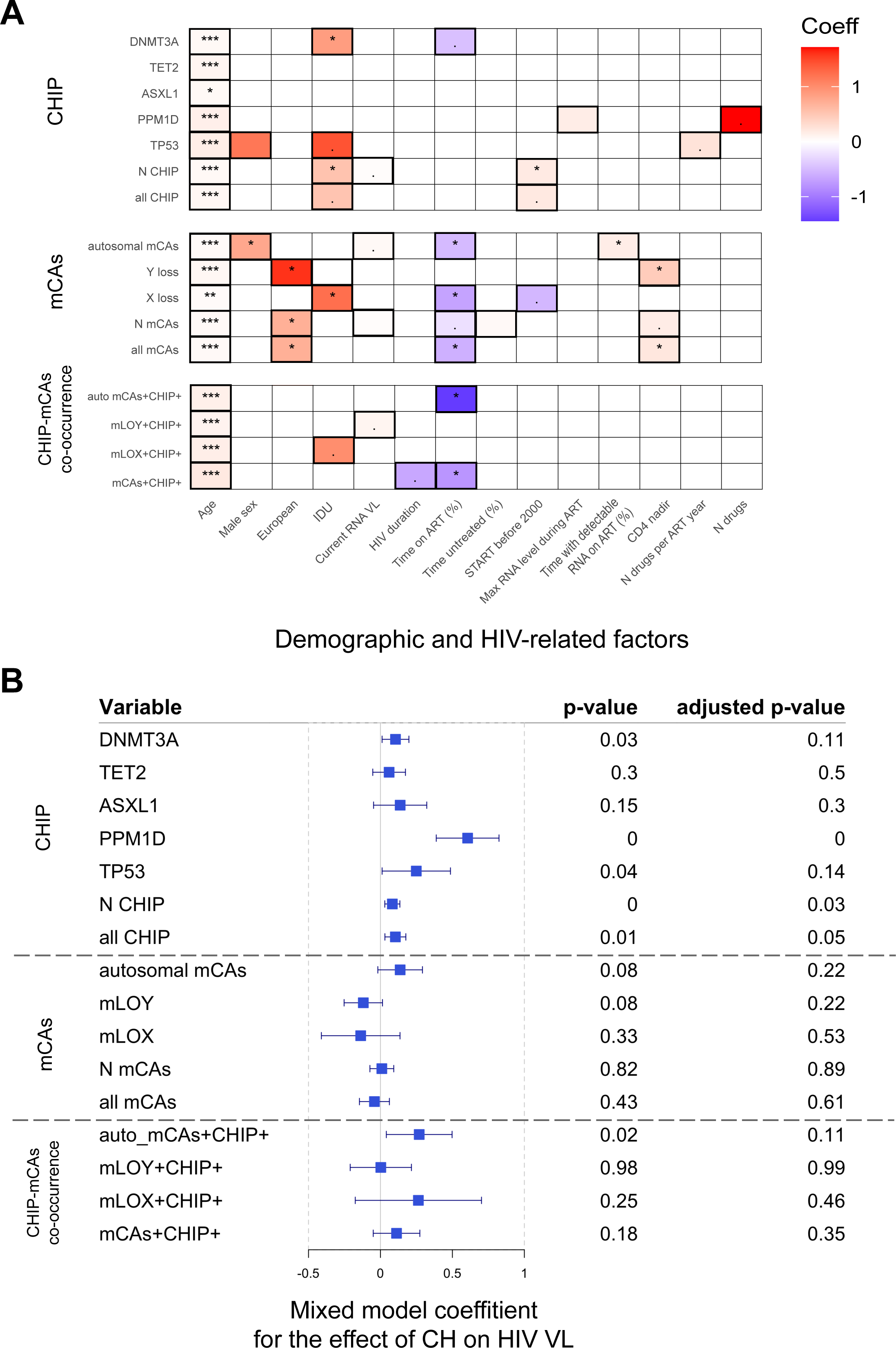
Association of demographic and HIV-related factors with different types of CH. A - Heatmap of regression coefficients from multivariable models. For each CH type (rows), a separate multiple regression model was fit: logistic regression for binary CH variables, and Poisson regression for the number of CHIP variants (N CHIP) and the number of mCAs (N mCAs). Variables retained after backward selection by at least one model are shown. Variables included to each model are highlighted and colored by the values of beta coefficients. White cells represent variables not included in the final model. Coeff - beta coefficient from logistic or Poisson regression. Different p-value thresholds represent the strength of the association:. p-value<0.1; * p-value<0.05; ** p-value < 0.01; *** p-value<0.001. IDU - injection drug use; Current RNA VL - mean HIV RNA viral load one year before and after CH identification; Time on ART (%) - the proportion of total HIV infection time during which the individual was receiving ART; Time untreated - proportion of the time of infection that was untreated; Start before 2000 - if the ART started before the year of 2000 (the proxy of treatment toxicity); Max RNA level during ART - maximum value of HIV RNA viral load during ART; Proportion detectable RNA ART - Proportion of the treated infection with detectable HIV RNA load; N drugs per ART year - total number of ART drugs taken normalized by the duration of ART; N drugs - total number of ART drugs taken. See Methods for the complete list of variables tested. B - Forest plot of mixed-effects models fitting CH status to longitudinal HIV RNA VL measurements (≥6 months after ART initiation). For each CH type, a separate mixed linear model was fit, adjusting for age, time since ART start, sex, ethnicity, smoking, and IVDU, with a random intercept for each individual. Squares represent model coefficients of CH effect on HIV RNA VL, and whiskers show 95% confidence intervals. A vertical dashed line at 0 indicates no association.

Some CH types (autosomal mCAs, number of CHIP variants, co-occurrence of CHIP and mLOY) were associated with higher HIV VL. To investigate if this relation is consistent throughout the course of HIV infection, we also performed a mixed linear model analysis for all measurements of HIV VL after ART initiation. We adjusted the model for age at blood sampling, duration of ART, sex, ethnicity, smoking, and IDU status. This analysis confirmed the association between higher HIV VL and CHIP but not mCAs (Figure 3B).

We also performed an exploratory analysis of the association of CH with different ART regimens and didn’t find any strong associations (Data not shown).

### Associations with blood counts

We investigated the potential effect of CH on specific blood count parameters, measured every 6 months, which reflect an inflammatory state or immune dysregulation. For association testing, we used two approaches: 1) a linear model of the mean of blood parameters one year before and after CHIP or mCAs screening versus CHIP/mCAs status, adjusting for age, sex, ancestry, smoking, and IDU status (Figure S1); 2) a mixed linear model of all measurements of blood parameters versus CHIP/mCAs status, adjusting for age, time since ART start, sex, ancestry, smoking, and IDU status, with random intercept for each individual (Figure 4). Both analyses yielded similar results, with more significant findings in the mixed model analyses: all CH types were associated with a lower CD4:CD8 ratio, mainly driven by higher CD8 T cell counts, whereas mCAs were associated with higher leukocyte, lymphocyte, and CD3 T cell counts (Figure 4).

**Figure 4.**
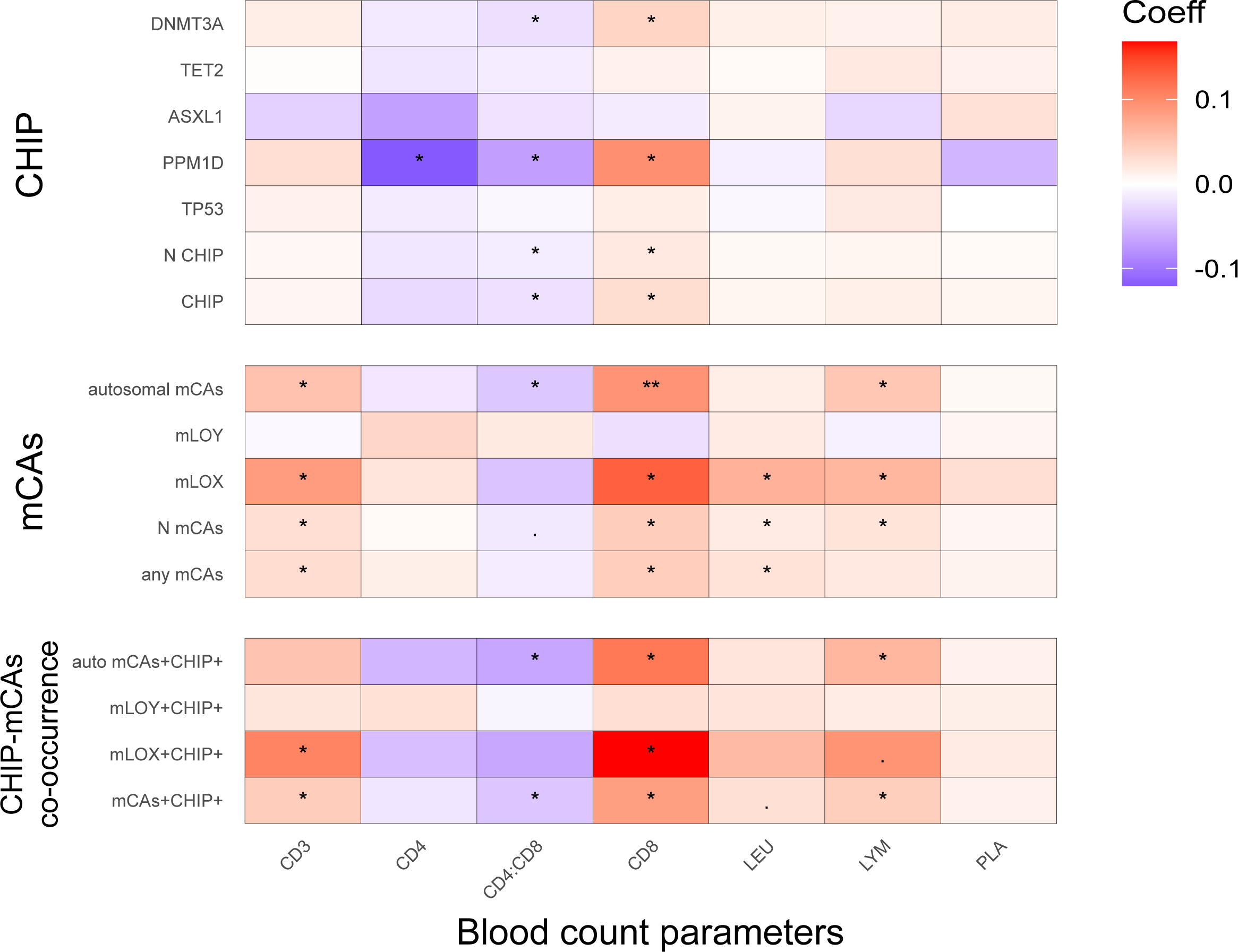
Associations of clonal hematopoiesis (CH) with longitudinal blood count parameters. Heatmap shows regression coefficients (color scale) from mixed linear models testing associations between each CH type (rows) and blood count parameters (columns), using measurements ≥6 months after ART initiation. For each blood feature and CH type, a separate mixed linear model was fit, adjusting for age, time since ART initiation, sex, ethnicity, smoking, and IDU, with a random intercept for each individual. Coeff - beta coefficient from mixed linear model for the effect of CH on blood features. Different p-value thresholds represent the strength of the association:. - p-value<0.1; * - p-value<0.05; ** - p-value < 0.01; *** - p-value<0.001. CD3 - CD3 lymphocyte count; CD4 - CD4 lymphocyte count; CD8 - CD8 lymphocyte count; CD4:CD8 - ratio of CD4 to CD8 lymphocyte count; LEU - leukocyte count; LYM - lymphocyte count; PLA - platelet count.

### Associations with hematological malignancies and aging-related diseases

To evaluate the clinical impact of CH in PWH on all-cause mortality and aging-related diseases, we conducted a survival analysis using the Cox proportional hazard model.

The majority of the CH types were associated with all-cause mortality (Figure 5). As expected, the presence of CHIP variants was associated with an increased risk of hematological and myeloid cancers; the presence of mCAs was not associated with an increased risk of any type of hematological malignancies, although their co-occurrence with CHIP increased the risk of lymphoid cancer. The number of CHIP variants and the presence of mLOX (especially when co-occurring with CHIP) were associated with the risk of solid tumors. mLOX and the presence of CHIP at a higher VAF (>10%, Supplementary Table 3) were associated with liver fibrosis and osteoporosis. mLOX was negatively associated with CKD, and no significant associations were found for CVD (Figure 5A; see Figure S2 for a larger set of CH types tested).

**Figure 5.**
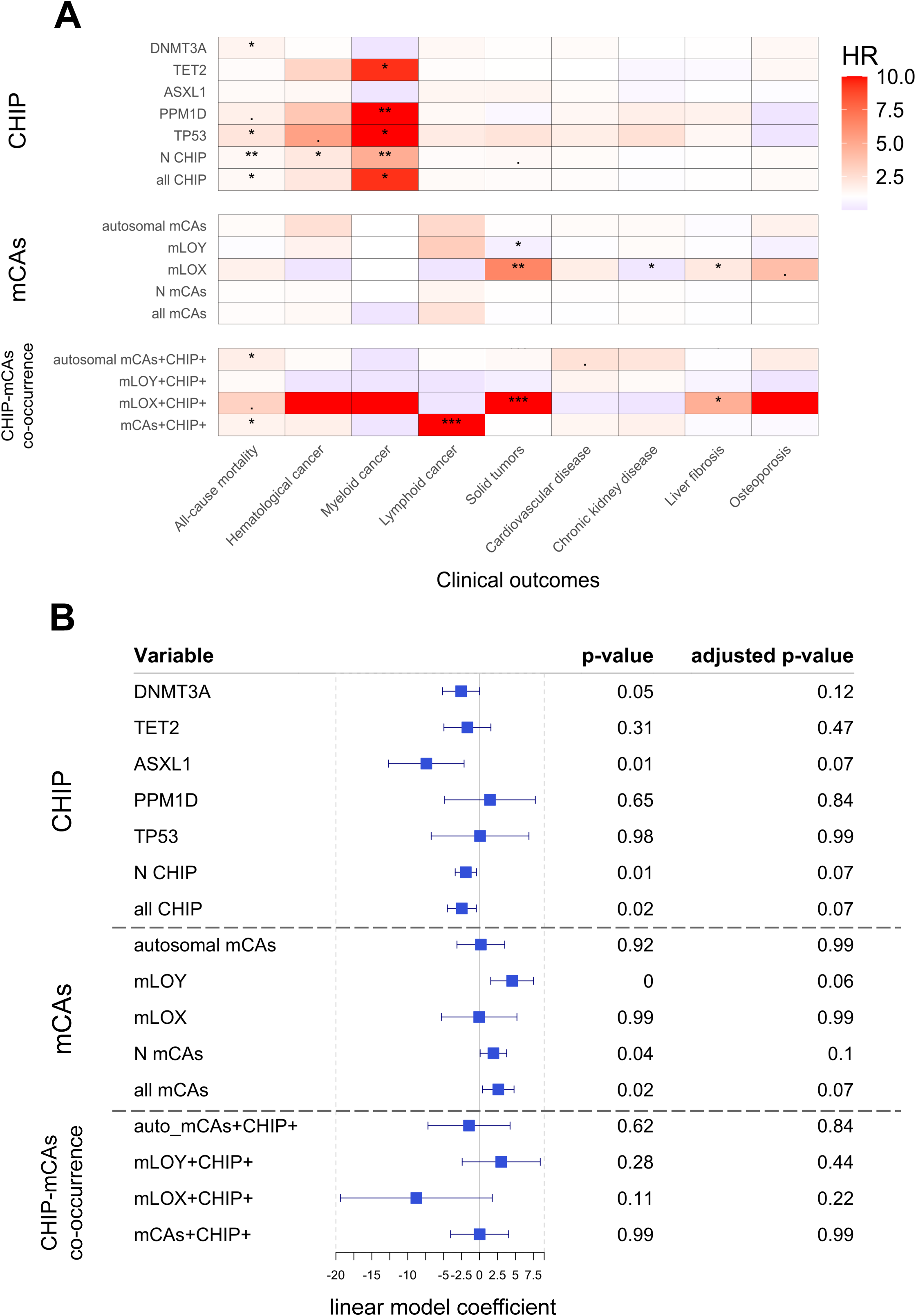
Associations of clonal hematopoiesis (CH) with clinical outcomes. A - Heatmap shows hazard ratios (HR, color scale) from Cox Proportional Hazard models testing associations between each CH type (rows) and clinical outcomes (columns). Separate models were fit for each disease and CH type and adjusted for a disease-specific set of covariates (see Methods). B - Forest plot of the results of the linear model showing the association of different CH types with the mean estimated glomerular filtration rate value within one year of CH screening. Each model was adjusted for the same set of covariates as for Chronic kidney disease. Different p-value thresholds represent the strength of the association:. - p-value<0.1; * - p-value<0.05; ** - p-value < 0.01; *** - p-value<0.001.

Although most CH types were not associated with CKD, CHIP was associated with lower estimated Glomerular Filtration Rate (eGFR) in the adjusted linear model, reflecting kidney function decline. mCAs were associated with higher eGFR, mostly driven by mLOY (Figure 5B).

## DISCUSSION

In this study, we assessed the prevalence and clinical associations of the two types of somatic variants causing clonal hematopoiesis - single-nucleotide variants in leukemoid genes (CHIP) and larger-scale genomic alterations (mCAs) - in a well-characterized population of PWH. We demonstrated that CH is both common and clinically relevant. One in four participants carried a CHIP variant, with a prevalence reaching 65% in individuals older than 85, and one in five harbored a mosaic chromosomal alteration, with prevalence rising sharply after age 65 to reach 60% (15% for autosomal mCAs) in individuals older than 85. The CHIP burden we observed exceeds the CHIP burden reported in population-based cohorts of similar age by approximately 10 years, and replicates earlier data ^22,23^, confirming that HIV infection and related factors (e.g., ART, comorbidities) predispose to somatic mutagenesis and/or clonal expansion of hematopoietic stem cells carrying CHIP variants. In contrast, the prevalence of mCAs was not higher than population expectations^6,17,34^, suggesting distinct mechanisms for point mutations versus chromosomal-scale somatic events leading to clonal expansion.

Age was the dominant risk factor for every type of CH, confirming a large number of previous reports. Conversely, neither CHIP nor mCAs correlated with smoking status. Such associations were reported repeatedly in the general population, but less consistently in PWH^23,35^. This suggests that traditional mutagens, like smoking, may play a lesser role in PWH, whose hematopoietic stem cells are subjected to other potential genotoxic influences like treatment-related toxicity or chronic immune activation. An additional association with CHIP was observed for IDU status, suggesting a potential mutagenic impact of repeated bouts of inflammation caused by injections, infections, or tissue trauma.

Among HIV–related factors, higher VL, a longer untreated infection (reflected by a lower proportion of time on ART relative to total infection duration), ART toxicity (approximated by initiation before the year 2000), and the number of antiretroviral drugs used were positively associated with several CH types (Figure 3). Toxicity of the first generation of ART and residual VL^35^ were previously proposed as risk factors for CHIP, but the supporting evidence was still inconclusive^23,24,35^. Our results confirm the potential deleteriousness of these factors, which can be minimized by the early prescription of modern, non-genotoxic ART.

Lower CD4:CD8 ratios were observed in CH carriers (Figure 4), possibly suggesting a positive feedback loop between immune dysfunction and clonal expansion. Persistently low CD4:CD8 ratios and elevated CD8 T cell counts, despite ART, represent an immunologic profile indicative of chronic activation and systemic inflammation^36,37^. Long-standing, low-grade inflammation may create a permissive environment for CH development. In turn, CH clones—particularly those harboring *TET2* or *DNMT3A* mutations—have been shown to amplify inflammation^11,24,38^, reinforcing this feedback loop in which HIV-driven immune activation promotes CH, and CH further sustains inflammation and hematopoietic dysfunction. The association between higher leukocyte count and mCAs also highlights a potential role of inflammation in the pathogenesis of CH. This cycle supports a model in which CH both reflects and accelerates immune aging, even during treated HIV infection. This could also explain the higher risk of severe inflammatory syndromes, such as immune reconstitution inflammatory syndrome (IRIS) and secondary hemophagocytic lymphohistiocytosis in PWH with CH^35,39^.

CH has emerged as a powerful biomarker of biological aging and complex disease risk in the general population. Numerous large-scale studies have demonstrated that individuals carrying CHIP or mCAs have an elevated risk of all-cause mortality, hematologic malignancies, and aging-related diseases such as cardiovascular disease and stroke^1,3,7,40^. In our cohort of PWH, we confirmed that most CH types are associated with increased all-cause mortality, independently of their strong association with age. However, we did not replicate some of the well-established links observed in the general population, such as associations between CH and cardiovascular or chronic kidney diseases. This may reflect limited statistical power due to the smaller cohort size and relatively low event counts for specific conditions, but it may also indicate differential baseline risk architecture in PWH. Chronic immune activation, metabolic disturbances, coinfections, and long-term antiretroviral exposure contribute substantially to cardiometabolic and inflammatory disease risk, possibly masking a more limited impact of CH. With regards to hematological cancers, CHIP predicted myeloid malignancies, as reported in the general population, but the presence of mCAs did not associate with lymphoid malignancies (although it showed a trend towards an increased risk), except when they co-occurred with CHIP^40^.

The enrichment in the co-occurrence of CHIP and autosomal mCAs (OR = 1.7) suggests that shared etiologies and/or selective pressures are involved in the generation of these CH types. Conversely, the absence of any enrichment in the co-occurrence of CHIP and sex chromosome losses suggests distinct mutational or selection mechanisms (e.g., mis-segregation errors, impact of inflammation) driving mLOY/mLOX.

To our knowledge, this is the first study to comprehensively assess the prevalence and clinical implications of sex chromosome loss — mLOY in men and mLOX in women — in PWH. While mLOY did not show significant associations with adverse outcomes in our study, in contrast to previous results from the general population^41,42^, multiple clinical correlations were found for mLOX. Specifically, mLOX was associated with an increased risk of solid tumors, liver fibrosis, and osteoporosis, as well as a decreased risk of chronic kidney disease (Figure 5), as opposed to the general population, where it was mostly associated with increased risk of leukemia^34^. These findings suggest that mLOX may have unique biological and clinical relevance among women with HIV and warrant further investigation into its pathophysiological mechanisms and prognostic value in this population.

Limitations of the current study include the cross-sectional nature of CH calling, which prevents the evaluation of clone dynamics; the use of a targeted sequencing panel restricted to 24 genes known to be involved in CHIP; the potential for survivor bias due to our selection of older study participants (≥65 years old) for the CHIP analyses; the different age distribution of the CHIP and mCA subpopulations included in our analyses; and limited event counts for some stratified analyses. Additionally, our study population was predominantly of European ancestry and from a single high-income country (Switzerland). This limits generalizability, as CH prevalence and patterns have been shown to vary by ancestry and environmental context^24^.

Given mounting evidence that CH is associated with accelerated biological aging, routine screening of high-risk PWH could be considered, especially as low-cost sequencing panels become feasible^26,40^. Interventions that have the theoretical potential to limit somatic mutagenesis or clonal expansion – like targeted anti-inflammatory treatments or minimization of ART toxicity – should be evaluated. Future work should include longitudinal evaluation of CHIP and mCAs to map clone appearance and trajectories, which is required to address the question of whether the elevated CHIP prevalence seen in PWH arises mainly from mutagenesis and clonal expansion during early untreated HIV infection, or from persistent inflammation and immune dysregulation even during long-term ART. Finally, it will be important to prospectively assess the potential benefit of early CH detection to better predict and prevent hematologic malignancies and complex aging-related conditions.

## Data Availability

The SHCS data are available to researchers upon the project submission and review by the Scientific Board of the SHCS and the study team; the provision of data is subject to Swiss legal and ethical regulations.

## Acknowledgements

This study has been financed within the framework of the Swiss HIV Cohort Study, supported by the Swiss National Science Foundation (grant #33FI-0_229621), by SHCS project #876, and by the SHCS research foundation. The data are gathered by the five Swiss University Hospitals, two Cantonal Hospitals, affiliated hospitals and private physicians.

PS’s institution has received travel grants, congress, and advisory fees from ViiV and Gilead unrelated to this work.

## Members of the Swiss HIV Cohort Study

Abela IA, Aebi-Popp K, Anagnostopoulos A, Bernasconi E, Braun DL, Bucher HC, Calmy A, Cavassini M (Chairman of the Clinical and Laboratory Committee), Ciuffi A, Dollenmaier G, Egger M, Elzi L, Fehr JS, Fellay J, Frigerio Malossa S, Fux CA, Günthard HF, Hachfeld A, Haerry DHU (deputy of “Positive Council”), Hasse B, Hirsch HH, Hoffmann M, Hösli I, Huber M, Jackson-Perry D (patient representatives), Kahlert CR, Kaufmann D, Keiser O, Klimkait T, Kouyos RD, Kovari H, Kusejko K (Head of Data Centre), Labhardt ND, Leuzinger K, Martinez de Tejada B, Marzolini C, Metzner KJ, Müller N, Nemeth J, Nicca D, Notter J, Paioni P (Chairman of the Mother & Child Substudy), Pantaleo G, Perreau M, Rauch A (President of the SHCS), Salazar-Vizcaya LP, Schmid P, Segeral O, Speck RF, Stöckle M, Surial B, Tarr PE, Trkola A, Wandeler G (Chairman of the Scientific Board), Weisser M, Yerly S.

## Authorship Contributions

AGB performed targeted CHIP sequencing and CHIP variants annotation. AC, MC, GC, HFG, LNW, PS, and PET contributed samples and data.

VT, KP, MAO, CWT, and JF performed statistical analysis and results interpretation. All authors contributed to the study design and preparation of the manuscript.

