## Supplementary material for "Epidemiology and Clinical Impact of Clonal Hematopoiesis in People with HIV": Figure S

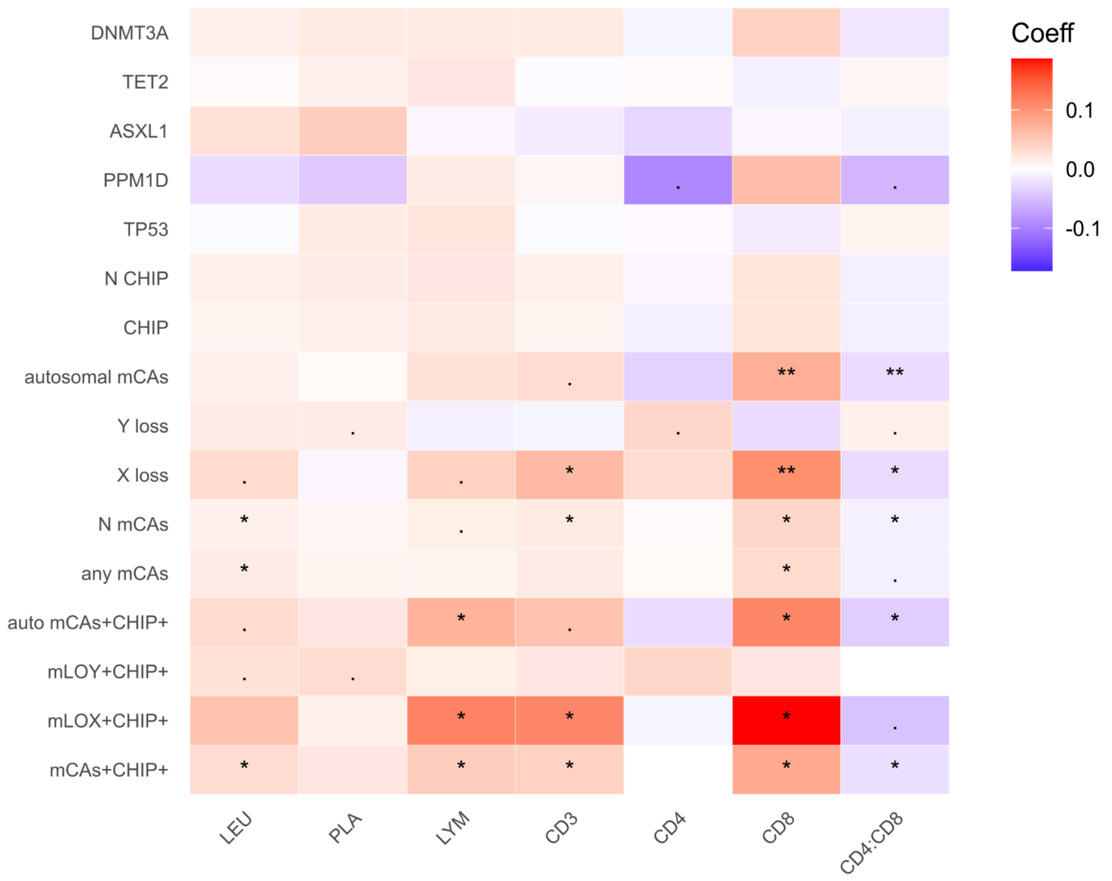


Figure S1. Associations of clonal hematopoiesis (CH) with cross-sectional blood count parameters calculated as the mean of blood count measurements within one year of CH screening. The heatmap displays regression coefficients (color scale) from linear models examining the associations between each CH type (rows) and blood count parameters (columns). For each blood feature and CH type, a separate linear model was fit, adjusting for age, time since ART initiation, sex, ethnicity, smoking, and IDU. Coeff - beta coefficient from linear model for the effect of CH on blood features. Different p-value thresholds represent the strength of the association: . - p-value<0.1; * - p-value<0.05; ** - p-value < 0.01; *** - p-value<0.001. CD3 - CD3 lymphocyte count; CD4 - CD4 lymphocyte count; CD8 - CD8 lymphocyte count; CD4:CD8 - ratio of CD4 to CD8 lymphocyte count; LEU - leukocyte count; LYM - lymphocyte count; PLA - platelet count.

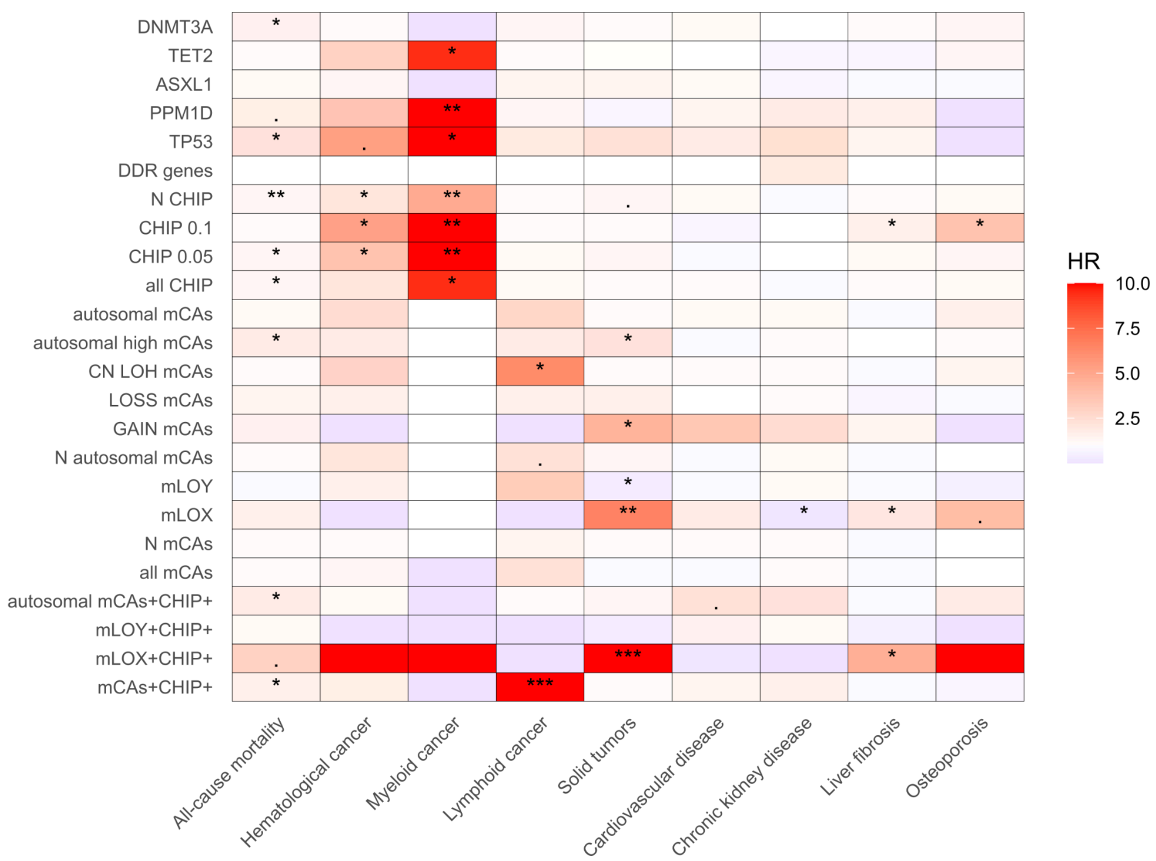


Figure S2. Associations of clonal hematopoiesis (CH) with clinical outcomes. This is an extended version of Figure 5, which includes additional CH types. Heatmap shows hazard ratios (HR, color scale) from Cox Proportional Hazard models testing associations between each CH type (rows) and clinical outcomes (columns). Separate models were fit for each disease and CH type and adjusted for a disease-specific set of covariates (see Methods). Different p-value thresholds represent the strength of the association: . - p-value<0.1; * - p-value<0.05; ** - p-value < 0.01; *** - p-value<0.001.
